# Validity of reported post-acute health outcomes in children with SARS-CoV-2 infection: a systematic review

**DOI:** 10.1101/2022.03.18.22272582

**Authors:** Julian Hirt, Perrine Janiaud, Viktoria Gloy, Stefan Schandelmaier, Tiago V. Pereira, Despina G. Contopoulos-Ioannidis, Steven N. Goodman, John P. A. Ioannidis, Klaus Munkholm, Lars G. Hemkens

## Abstract

**Importance:** There is concern that post-acute SARS-CoV-2 infection health outcomes (“post-COVID syndrome”) in children could be a serious problem but at the same time there is concern about the validity of reported associations between infection and long-term outcomes.

**Objective:** To systematically assess the validity of reported post-acute SARS-CoV-2 infection health outcomes in children.

**Evidence Review:** A search on PubMed and Web of Science was conducted to identify studies published up to January 22, 2022, that reported on post-acute SARS-CoV-2 infection health outcomes in children (<18 years) with follow-up of ≥2 months since detection of infection or ≥1 month since recovery from acute illness. We assessed the consideration of confounding bias and causality, and the risk of bias.

**Findings:** 21 studies including 81,896 children reported up to 97 symptoms with follow-up periods of 2-11.5 months. Fifteen studies had no control group. The reported proportion of children with post-COVID syndrome was between 0% and 66.5% in children with SARS-CoV-2 infection (n=16,986) and 2% to 53.3% in children without SARS-CoV-2 infection (n=64,910). Only 2 studies made a clear causal interpretation of an association of SARS-CoV-2 infection and the main outcome of “post-COVID syndrome” and provided recommendations regarding prevention measures. Two studies mentioned potential limitations in the conclusion of the main text but none of the 21 studies mentioned any limitations in the abstract nor made a clear statement for cautious interpretation. The validity of all 21 studies was seriously limited due to an overall critical risk of bias (critical risk for confounding bias [n=21]; serious or critical risk for selection bias [n=19]; serious risk for misclassification bias [n=3], for bias due to missing data [n=14] and for outcome measurement [n=12]; and critical risk for selective reporting bias [n=16]).

**Conclusions and Relevance:** The validity of reported post-acute SARS-CoV-2 infection health outcomes in children is critically limited. None of the studies provided evidence with reasonable certainty on whether SARS-CoV-2 infection has an impact on post-acute health outcomes, let alone to what extent. Children and their families urgently need much more reliable and methodologically robust evidence to address their concerns and improve care.

**KEY POINTS:** *Question:* How valid are the reported results on health outcomes in children after acute SARS-CoV-2 infection?

*Findings:* We identified 21 studies with only 6 using a controlled design. Reported post-acute health-outcomes were numerous and heterogeneous. The reported proportion of children with post-COVID syndrome was up to 66.5% in children with and 53.3% in children without SARS-CoV-2 infection. All studies had seriously limited validity due to critical and serious risk of bias in multiple domains.

*Meaning:* The validity of reported post-acute SARS-CoV-2 infection health outcomes in children is critically limited and methodological robust evidence is urgently needed.

## INTRODUCTION

Children usually have no or mild symptoms from the severe acute respiratory syndrome coronavirus 2 (SARS-CoV-2) infection^1,2^ and are rarely hospitalized with extremely rare fatal events^3^. However, symptoms persisting beyond the acute stage have been observed in adults^4^, but also in children^3^. Such persistent symptoms or post-acute health outcomes are often referred to as long-COVID or post-COVID syndrome, but there is no consensus on how to define it^5^. The term long-COVID may encompass both ongoing symptomatic COVID-19 (4-12 weeks after the initial infection) and post-COVID-19 syndrome (>=12 weeks after the initial infection)^6^ or defined as post-COVID-19 condition occurring 3 months from the onset of COVID-19 with symptoms that last for at least 2 months^7^.

Evidence syntheses aiming to assess post-acute SARS-CoV-2 infection health outcomes in children are difficult given the highly heterogenic study designs and complex limitations. There are methodological concerns about the validity of reported causal effects of infection on long-term outcomes^3,4^, including the absence of a control group, missing outcome data, and detection and misclassification biases^4^. Moreover, there is serious heterogeneity in study populations (diverse settings, eligibility criteria, and sampling strategies) and lack of standardization of cases and outcomes^3,4^ that limits the interpretation of estimates for absolute risks and post-covid syndrome prevalence. The validity of relative risk estimates critically depends on the risk for confounding bias resulting from systematic differences between the compared groups. However, confounding is often ignored when interpreting epidemiologic studies^8,9^, and their authors rarely call for cautious interpretation^9^.

Given the minimal risk of serious acute outcomes, post-acute health outcomes may be the actual critical child health problem caused by SARS-CoV-2 infection^10-12^. Valid estimates of this risk are crucial for valid assessments of benefits and harms of preventive measures in children, including nonpharmaceutical interventions (e.g., masks or school closures) and vaccines. Given the potential impact on child health worldwide, we would expect that studies assessing these risks are conducted with the greatest possible care and the intention to meet highest available standards in reporting and transparency to deal with confounding, other biases, and causal claims.

Here, we systematically assessed the validity of reported outcomes of studies that aimed to determine the effect of SARS-CoV-2 infection on post-COVID syndrome in children.

## METHODS

We systematically identified studies reporting post-acute health outcomes in children with SARS-CoV-2 infections using a similar search strategy and study selection as a previous related analysis^10^. Our study protocol is published^13^ and no major deviations occurred. We used the “Preferred Reporting Items for Systematic reviews and Meta-Analyses 2020” (PRISMA 2020)^14^. Our analysis was used to expand previous methodological recommendations^10^ to increase the validity of future research on post-COVID syndrome in children.

### Eligibility criteria

Eligible studies included a cohort of children (<18 years) defined by the presence of SARS-CoV-2 infection; reported frequency of health outcomes (i.e., any symptoms) for this cohort of children (accepting subgroup analyses); had a clearly defined follow-up of ≥ 2 months since detection of SARS-CoV-2 infection, onset of symptoms, COVID-19 diagnosis and/or hospital admission, or a clearly defined follow-up ≥ 1 month since recovery from acute illness and/or hospital discharge (follow-up defined as in previous related analysis^10^); and were published as preprint or peer-reviewed journal article in English.

We considered studies regardless of the severity of the acute SARS-CoV-2 infection or the setting (e.g., outpatient, hospitalized, intensive care unit), and regardless of whether the investigation included a comparator group of participants without SARS-CoV-2 infection. We excluded meta-analyses and evidence-syntheses, abstract-only publications, and conference proceedings.

### Information sources and search strategy

We adapted an existing search strategy^4^ (Supplementary material: eMethods) and searched PubMed and Web of Science Core Collection since January 1, 2020; the living systematic COVID-19 map provided by the EPPI-Centre (using keywords for the pediatric population^15^), and the Living Overview of Evidence platform for COVID-19 (L·OVE) platform (to identify preprints)^16^. Date of the last search was January 22 (for PubMed and Web of Science Core Collection) and January 25 (for L·OVE and EPPI-Centre), 2022. We contacted investigators of registered systematic reviews on long-COVID in children to cross-check eligible studies^17-19^ (last PROSPERO search November 5, 2021; authors’ request November 23, 2021); and also screened citations of further relevant reviews^3,20,21^.

### Study selection

One researcher screened titles and abstracts (JH, PJ, or LGH). Potentially relevant full texts were screened by two researchers (two of JH, PJ, StS, TVP, or VG) independently. Any disagreements were resolved by discussion or third-party arbitration (LGH).

### Data extraction and methodological appraisal

We extracted study characteristics (e.g., study design, population, exposure, outcomes, results) and assessed the consideration of confounding bias and causality, and the risk of bias^8,9,22,23^. Details on extracted items and algorithms for assessments were prespecified^13^. Extractions and assessments were performed by two independent researchers (two of JH, PJ, StS, TVP, or VG). Disagreements were resolved by discussion or with a third reviewer (LGH).

#### Targeted outcomes of eligible studies

We considered symptoms and quality of life outcomes or other patient-relevant outcomes reflecting how children feel, function (or survive)^24^. Imaging or laboratory measures were not considered. We determined the most frequent symptoms assessed among all studies. We recorded the number of reported and total assessed (i.e., as reported in the protocol, registry, or methods section) outcomes; source of outcome definition (e.g., expert opinion, literature-based questionnaire); outcome data collection method (e.g., phone, online, clinical visit); length of follow-up (median or mean, as reported; in studies where only a range was reported, we used the midpoint); reported duration of symptoms (i.e., persistent, episodic, or single event); reported frequency of symptoms (e.g., several times daily, once per day or week); reported trend of persistent symptoms (i.e., improving or worsening); reported severity of symptoms. We recorded the proportions of children presenting the main outcome. The main outcome was defined as stated by the authors or, if not available, the most inclusive one (e.g., any symptoms)^13^. We also assessed if the main outcome was analyzed in relation to specific participant characteristics (e.g., comorbidities) or infection-related factors (e.g., severity acute disease).

#### Consideration of confounding bias and causality, and risk of bias assessment

We assessed the consideration of confounding using a previously developed approach^8,9^, based on prespecified questions focusing on the reporting of confounders and bias in the abstract and discussion, and on what the findings mean and what the limitations are.

We assessed three aspects related to consideration of causality, following a similar but simplified approach as Haber et al.^22^. We determined whether an association of SARS-Cov-2 infection and the main outcome was interpreted causally and if any recommendations were made based on such causal implications; whether a conceptual causal model describing causal mechanisms (e.g., a directed acyclic graph^25,26^) had been used; and whether any explicit causal disclaimer was made. We assessed the risk of bias for an estimated effect of SARS-Cov-2 infection on the main post-acute symptom outcome using “Risk Of Bias In Non-randomised Studies – of Interventions” (ROBINS-I)^23,27^. We adapted the tool specifically to this non-interventional situation^13^.

### Statistical analysis

We used R (version 4.1) for all analyses. We report medians and interquartile ranges (IQR), and calculated proportions with 95% confidence intervals (CIs) using the “metaprop” function from the “meta” package (version 5.1-1^28^) in R.

## RESULTS

Twenty-one studies were eligible (Table 1; Supplementary material: eFigure1). Six studies had a control group^29-34^ and 15 were uncontrolled^35-49^. Twelve studies were conducted in Europe ^29-33,36,38-40,44,47,49^ and 18 published in 2021^29-33,35-37,39-48^. Thirteen studies used data that were specifically collected for the purpose of the study^29,31-33,35-40,42,45,47,48^, 5 studies used routinely collected data^30,34,41,44,49^ and the type of data was unclear in 2 studies^43,46^. Only 2 studies were registered^30,32^. Availability of study protocol was mentioned in 3 studies^31,32,42^ with one of them not publicly available^42^. Nineteen studies reported ethical approval, 1 did not report on this^38^, and 1 did not undergo full ethical review^44^.

**Table 1:** Characteristics of cohort studies reporting post-acute health outcomes in children with SARS-CoV-2 infection.

|  | Total<br>No. (%) | Controlled<br>No. (%) | Uncontrolled<br>No. (%) |
| --- | --- | --- | --- |
| **Total** | 21 (100) | 6 (28.6) | 15 (71.4) |
| **Region** |  |  |  |
| Europe | 12 (57.1) | 5 (83.3) | 7 (46.7) |
| Asia* | 6 (28.6) | 0 (0) | 6 (40) |
| North America | 2 (9.5) | 1 (16.7) | 1 (6.7) |
| Oceania | 1 (4.8) | 0 (0) | 1 (6.7) |
| **Population** |  |  |  |
| Hospitalized only | 12 (57.1) | 1 (16.7) | 11 (73.3) |
| Hospitalized and outpatient | 4 (19.0) | 1 (16.7) | 3 (20) |
| General population | 4 (19.0) | 4 (66.7) | 0 (0) |
| Emergency department only | 1 (4.8) | 0 (0) | 1 (6.7) |
| **Sample size** |  |  |  |
| <100 | 12 (57.1) | 1 (16.7) | 11 (73.3) |
| 100 - 500 | 4 (19.0) | 1 (16.7) | 3 (20.0) |
| 501 - 1000 | 2 (9.5) | 1 (16.7) | 1 (6.7) |
| >1000 | 3 (14.3) | 3 (50.0) | - |
| **Definition of exposure (SARS-CoV-2 infection)** |  |  |  |
| Using RT-PCR only | 11 (52.4) | 1 (16.7) | 10 (66.7) |
| Using RT-PCR or serology | 5 (23.8) | 2 (33.3) | 3 (20.0) |
| Using RT-PCR, antigen, or serology | 2 (9.5) | 1 (16.7) | 1 (6.7) |
| Unclear | 2 (9.5) | 1 (16.7) | 1 (6.7) |
| Using serology only | 1 (4.8) | 1 (16.7) | 0 (0) |
| **Definition of no-exposure (control group)** |  |  |  |
| No diagnosis or symptoms | - | 3 (50.0) | - |
| Using serology | - | 2 (33.3) | - |
| Using RT-PCR | - | 1 (16.7) | - |
| **Study registration** |  |  |  |
| Mentioned | 2 (9.5) | 2 (33.3) | 0 (0) |
| Not mentioned | 19 (90.5) | 4 (66.7) | 15 (100) |
| **Study protocol** |  |  |  |
| Mentioned | 2 (9.5) | 2 (33.3) | 0 (0) |
| Not mentioned | 19 (90.5) | 4 (66.7) | 15 (100) |
| **Ethical approval** |  |  |  |
| Yes | 19 (90.5) | 6 (100) | 13 (86.7) |
| No full review ** | 1 (4.8) | 0 (0) | 1 (6.7) |
| Unclear | 1 (4.8) | 0 (0) | 1 (6.7) |

\* Including Russia; \*\* "As the data analysis was retrospective and no additional data were collected beyond those required for standard medical care, a full ethics review under the terms of the Governance Arrangements of Research Ethics Committees in the UK was not required."<sup>44</sup>
Abbreviations: No. = Number.

### Study populations

Overall, the 21 studies included 81,896 children (range 14-71,700; median 58; IQR 25-151)^29-49^. The uncontrolled studies included 1,335 children with SARS-CoV-2 infection (range 14-518; median 50; IQR 19-106). The controlled studies included 15,651 children with SARS-CoV-2 infection (range 16-11,950; median 280; IQR 72-2,411) and 64,910 children without SARS-CoV-2 infection (range 17-59,750; median 672; IQR 69-3,115). Nine studies had a sample size of more than 100 children (including controls)^30-34,40,42,46,49^. Children were analyzed as a subgroup in 4 studies^29,30,37,41^. Four studies, all controlled, primarily recruited children from the general population (i.e., random sample from schools in Switzerland^31^; consecutive sample of home-isolated children in Norway^29^; based on large health insurance databases in Germany^30^; and based on a national health system database in the United Kingdom^32^). Seventeen studies recruited only hospitalized children^34-36,38,41-48^, only children attending the hospital as outpatient^39^, or both^33,37,40,49^. Five of these 17 studies included children with multisystem inflammatory syndrome (MIS-C)^34,43,44,48,49^.

Positive SARS-CoV-2 infection status was confirmed in 11 studies using RT-PCR only^32,35-42,45,47^, 7 using a mix of tests (RT-PCR, antigen test, or serology)^29,33,34,43,44,48,49^, 2 did not provide information on the test used^30,46^, and 1 used serology only^31^.

Negative infection status in the 6 controlled studies was defined in 3 studies as no diagnosis or symptoms of acute COVID-19^30,33,34^, in 2 by serology testing^29,31^, and in 1 study determined by RT-PCR^32^.

#### Outcome data collection methods and outcome ascertainment

Outcome data were collected over 2 to 7 months (median 5.3; IQR 3-6) in the 12 studies starting follow-up at infection or onset^29-34,39,44,46-49^ and over 2 to 11.5 months (median 5; IQR 3.5-8) in the 9 studies starting follow-up at recovery^35-38,40-43,45^ (Table 2; Supplementary material:). Outcome data were collected by phone in 8 studies^33,35,37,38,40,42,45,47^, per clinical visit in 5^36,39,43,46,48^, routinely collected in 4^30,34,44,49^, online in 2^30,31^, by personal interview in 1^29^, or by a mix of case reports, medical records, and self-reports in 1^40^.

**Table 2:** Details on reported post-acute health outcomes.

| Study | Follow-up (months) * | Main outcome | Number of outcomes (reported/total assessed) | Outcome data collection method | Symptom duration reported | Symptom frequency reported | Symptom trend reported | Symptom severity reported | Subgroup analyses related to main outcome |
| --- | --- | --- | --- | --- | --- | --- | --- | --- | --- |
| **Controlled studies** |  |  |  |  |  |  |  |  |  |
| Bergia 2021<sup>33</sup> | 4 (infection) | <u>any symptoms</u> | 20/n.r. | Structured questionnaire by phone led by physicians | Yes | - | - | - | Yes |
| Blomberg 2021<sup>29</sup> | 6 (infection) | <u>any symptoms</u> | 11/11 | Personal interview led by medical staff | - | - | - | - | - |
| Matsubara 2022<sup>34</sup> | 3 (infection) | <u>cardiac symptoms</u> | 3/n.r. | Routinely collected data (medical records, structured clinical assessment) | - | - | - | - | - |
| Radtke 2021<sup>31</sup> | 6 (infection) | <u>any symptoms</u> | 11/n.r. | Structured online questionnaire | Yes | - | - | - | - |
| Roessler 2021<sup>30</sup> | 3 (infection) | <u>health outcomes combined</u> | 97 (grouped)/97 | Routinely collected data (administrative claims; unclear how symptoms were assessed) | - | - | - | - | Yes |
| Stephenson 2021<sup>32</sup> | 3 (infection) | <u>any symptoms</u> | 25/25 | Structured online questionnaire | - | - | - | - | Yes |
| **Uncontrolled studies** |  |  |  |  |  |  |  |  |  |
| Asadi-Pooya 2021<sup>35</sup> | 8 (recovery) | <u>any symptoms</u> | 28/28 | Structured questionnaire by phone (unclear who led by) | - | Yes | - | Yes | Yes |
| Bottino 2021<sup>36</sup> | 2 (recovery) | <u>respiratory symptoms</u> | 1/n.r. | Clinical visit (unclear who led by and how symptoms were assessed) | - | - | - | - | - |
| Capone 2021<sup>48</sup> | 6 (infection) | <u>fatigue</u> | 1/n.r. | Clinical visit (unclear who led by and how symptoms were assessed) | - | - | - | - | - |
| Chowdhury 2021<sup>37</sup> | 5 (recovery) | <u>any symptoms</u> | 16/n.r. | Phone call (unclear who led by and which symptoms were assessed) | Yes | - | - | - | - |
| Denina 2020<sup>38</sup> | 4 (recovery) | <u>any symptoms</u> | 1/n.r. | Phone call (unclear who led by and which symptoms were assessed) | - | - | - | - | - |
| Isoldi 2021<sup>39</sup> | 6 (infection) | <u>any symptoms</u> | 1/n.r. | Clinical visit (unclear who led by and how symptoms were assessed) | Yes | - | - | - | - |
| Kahn 2022<sup>49</sup> | 2 (infection) | <u>any symptoms</u> | 9/n.r. | Routinely collected data (registry; structured clinical assessment) | - | - | - | - | - |
| Matteudi 2021<sup>40</sup> | 11.5 (recovery) | <u>any symptoms</u> | 6/n.r. | Phone call led by a paediatric team (unclear which symptoms were assessed) | - | - | Yes | - | Yes |
| Mei 2021<sup>41</sup> | 5 (recovery) | <u>any symptoms</u> | 1/48 | Case reports, medical records, self-reports (unclear who led by and how symptoms were assessed) | - | - | - | - | - |
| Osmanov 2021<sup>42</sup> | 8.5 (recovery) | <u>any symptoms</u> | 44/44 | Structured questionnaire by phone led by medical students | Yes | Yes | Yes | - | Yes |
| Patnaik 2021<sup>43</sup> | 3.5 (recovery) | <u>any symptoms</u> | 1/n.r. | Clinical visit (unclear who led by and how symptoms were assessed) | - | - | - | - | - |
| Penner 2021<sup>44</sup> | 6 (infection) | <u>gastrointestinal symptoms</u> | 9/9 | Routinely collected data (medical records; structured clinical assessment) | Yes | - | Yes | - | - |
| Rusetsky 2021<sup>45</sup> | 2 (recovery) | <u>olfactory disorder</u> | 1/1 | Structured questionnaire by phone led by the investigators | Yes | - | - | Yes | Yes |
| Say 2021<sup>46</sup> | 4.5 (infection) | <u>any symptoms</u> | 24/24 | Clinical visit using a structured clinical assessment (unclear who led by) | Yes | - | - | - | - |
| Sterky 2021<sup>47</sup> | 7 (infection) | <u>any symptoms</u> | 18/18 | Structured questionnaire by phone (unclear who conducted) | - | - | - | Yes | Yes |

\* Follow-up started at detection of infection, onset of symptoms, COVID-19 diagnosis and/or hospital admission (described as infection) or at recovery from the acute illness and/or hospital discharge (described as recovery).
Abbreviations: n.r. = not reported.

Eight studies considered data on symptom duration^31,33,37,39,42,44-46^. Symptom trend (n=3)^40,42,44^, severity (n=3)^35,45,47^, and frequency (n=2)^35,42^ were uncommonly considered. The main outcome was analyzed in relation to participant characteristics or infection-related factors in 8 studies^30,32,33,35,40,42,45,47^.

#### Outcome definition and outcome types

Only 2 studies provided clear background information on how they defined post-acute symptoms (i.e., questionnaire based on international working group^31^; expert opinion and published literature^30^; Table 2; Supplementary material: eTable1).

Thirteen studies focused only on symptoms^30-33,35,40-43,45-48^ while the others also reported numerous laboratory parameters^29,34,36-39,44,49^. Reported symptoms ranged from 1 to 97 per study (median 9; IQR 1-20), however, for 11 of the 21 studies the total number of assessed outcomes was unclear^31,33,34,36-40,43,48,49^.

The main outcome was a composite of any symptom in 16 studies^29-33,35,37-43,46,47,49^; the main outcome in the remaining studies was fatigue^48^, respiratory^36^, gastrointestinal^44^, cardiac symptoms^34^, and olfactory symptoms^45^. The 3 symptoms most frequently assessed across studies were headache (n=13)^29-33,35,37,40-42,46,47,49^, fatigue or tiredness (n=12)^30-32,34,35,37,41,42,46-49^, and cough (n=10)^29-33,35,37,41,42,46^.

#### Outcome results

The reported proportion of children with post-COVID syndrome (main post-acute health outcomes) was between 0% and 66.5% in children with SARS-CoV-2 infection (median 13%; IQR 0-22%; 17 studies; Figure 1). Between 2% and 53.3% of children without SARS-CoV-2 infection also had such symptoms (control groups of 6 studies; Figure 1).

**Figure 1:**
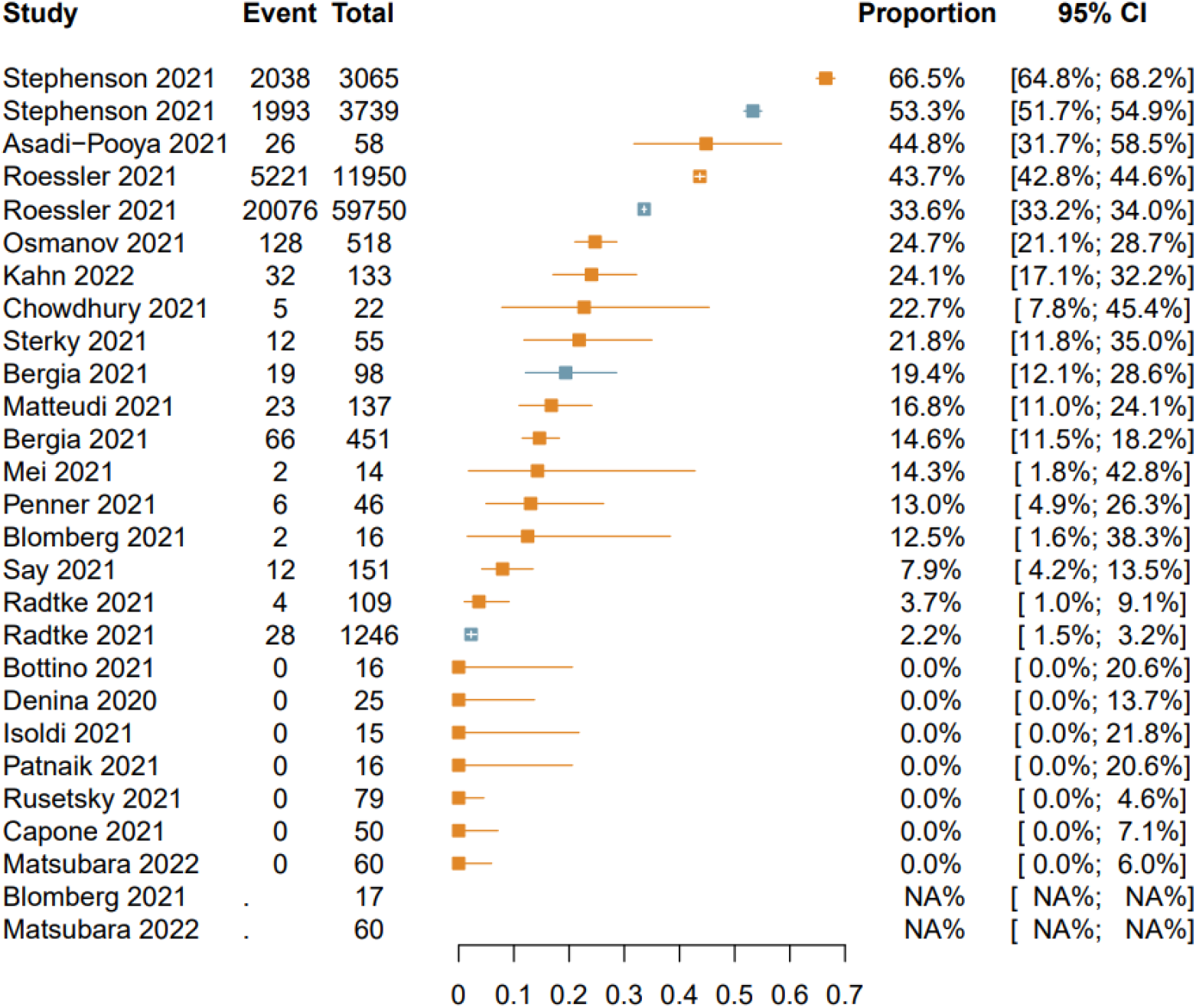
Proportions of children with reported main post-acute health outcomes. Orange-colored: estimates and 95%-CI for children with SARS-CoV-2 infection; blue-colored: estimates and confidence intervals for children without SARS-CoV-2 infection in the 6 controlled studies^29-34^. No data reported for children without SARS-CoV-2 infection Blomberg 2021^29^ and Matsubara 2022^34^. Abbreviations: CI = Confidence interval.

Only one study described a formal statistical comparison between SARS-Cov-2 infected children and controls, reporting an incidence rate ratio of 1.30 (95% CI 1.25 to 1.35) for all health outcomes combined^30^.

#### Consideration of confounding

Sixteen of the 21 studies did not allude to or mention confounding bias at all^29,31,35-48^. Three studies alluded to confounding bias^33,34^ or acknowledged specific non-adjusted confounders^34,49^, with one of them presenting a statement on residual confounding^33^.

Two studies statistically considered confounding bias^30,32^ (by matching on age, sex, geographical area^32^ and matching on age, sex, comorbitites^30^). Only one study clearly discussed the issue of confounding: *“We cannot exclude that our results may be affected by unmeasured confounding, although we minimized differences between COVID*□*19 and control cohort via matching”*^30^.

Ten studies clearly discussed other biases, for example as information, referral, detection, response, or recall bias, or alluded to other potential biases affecting, for example, missing data^30-35,42,44,48,49^. Potential limitations were mentioned in the conclusion of the main text of 2 studies^30,31^ while no study mentioned any limitations nor made a clear statement for cautious interpretation in the abstract.

#### Consideration of causality

A clear causal interpretation of an association between SARS-CoV-2 infection and the main outcome was made in 2 of the 21 studies, both were controlled studies and recommended actions regarding prevention measures^29,30^ (Supplementary material: eBox1). In only 1 of these 2 studies did the authors mention that their results may be impacted by confounders^30^. The same study was also the only study providing a clear causal disclaimer in the discussion stating *“Due to the observational nature of our study, a main limitation is that its design does not induce a causal interpretation of results”*^30^. No studies used a conceptual causal model.

#### Risk of bias

All 21 studies had an overall critical risk of bias with critical risk in at least one domain (Figure 2; Supplementary material: eTable2). Risk of bias due to confounding was critical in all studies^29-49^. This was because uncontrolled studies are unable ^35-49^, by design, to adjust for confounders and because controlled studies^29-34^ either did not provide a formal statistical comparisons between infected children and controls^29,31,33,34^ or did not adjust for relevant confounders^30,32^ such as family and housing settings, and socioeconomic status.

**Figure 2:**
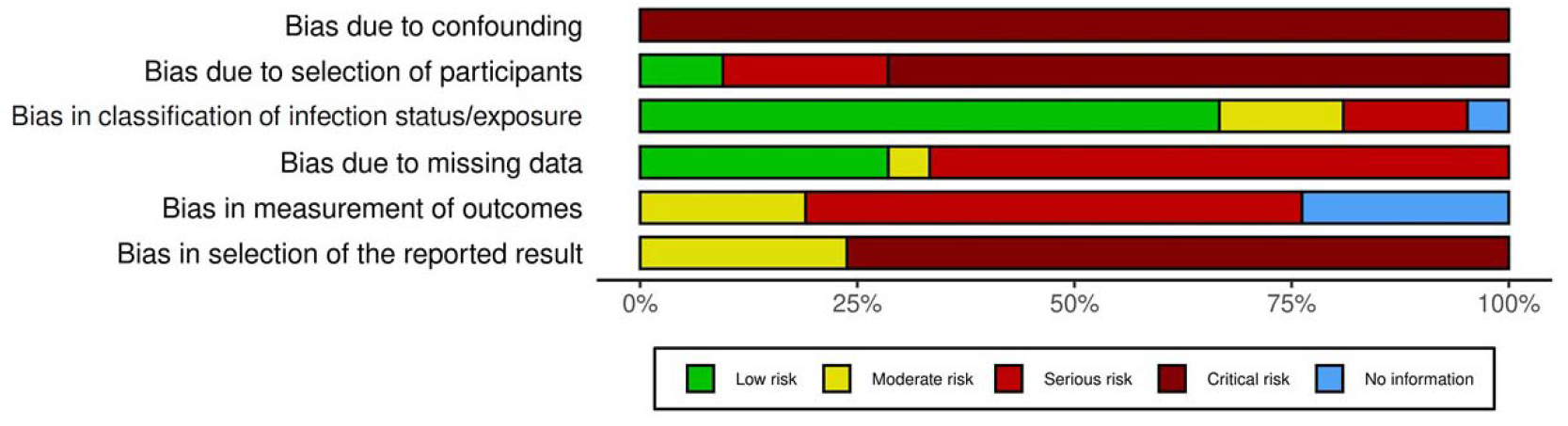
Summary of risk of bias assessment.

Risk of bias due to selection of participants was serious or critical in all studies except for 2 controlled studies that recruited from electronic health databases regardless of acute symptoms and hospitalization^30,32^. The risk of bias resulted mainly from self-selection of participants as they were aware of their infection status in all studies.

Risk of misclassification bias due to ambiguous definition of infection status was low for 14 uncontrolled studies that identified patients based on RT-PCR tests during acute COVID-19^30,32,35-44,47-49^.Three controlled studies^29,31,32^ had moderate risk of bias either because positive infection status was defined by serology, but the infection time point was unclear or because negative infection status was defined by RT-PCR, but previous infection could not be excluded. The other 3 controlled studies^30,33,34^ had serious risk of bias because negative infection status was determined on the basis of having no documented COVID-19 diagnosis^30^ or symptoms^33,34^ regardless of laboratory virus detection. Risk of bias due to missing data was serious in 14 studies, because loss-to-follow-up was either>20%^29,31-34,37,39,40,42,43,48,49^ or the information provided was unclear^35,38^. One study^41^ reported following all participants but used multiple data sources without providing further details and was deemed to be of moderate risk of bias. Studies with low risk of bias reported no or minimal loss-to-follow-up^36,45-47^, or used systematically routinely collected data^30,44^.

Risk of bias in measurement of outcomes was serious for 12 studies that used self-report methods^28-32,34,36,37,39,41,44,46^; we assumed that participants who knew they were infected were more likely to report symptoms. Risk of outcome measurement bias was moderate for 4 studies that used routinely collected data based on structured clinical assessment^34,44,49^ or assessed outcomes during a clinic visit using structured clinical assessment^46^.

Risk of bias in selection of the reported outcome was critical in 16 studies either because they did not provide a protocol and/or the questionnaire used, failed to clearly define outcomes, and/or did not report all outcomes assessed^29,31-35,37-41,43,44,46,48,49^.

## DISCUSSION

This systematic review of 21 studies found critically limited validity of reported SARS-CoV-2 infection health outcomes in children that may be perceived as post-COVID syndrome. There was huge heterogeneity in the definition, assessment, and reported frequency of symptoms with frequently missing important information. Overall, none of the studies provided evidence with reasonable certainty on whether SARS-CoV-2 infection has an impact on post-acute health outcomes, let alone to what extent.

Several ongoing systematic reviews aim to determine the prevalence of persistent of symptoms associated with SARS-CoV-2 infection in children^17-19^, and one has been published^20^. However, none specifically assessed the validity of reported post-acute symptoms with a focus on an integrative assessment of confounding, other biases, and causal claims in this emerging research field. We avoided any quantitative synthesis of risk estimates given the biases and heterogeneity in these studies; any combined estimate would most likely be misleading for patients, parents, clinicians, the general public, and policymakers. Children in the control groups of the two largest studies^30,32^ very often had symptoms of post-COVID syndrome without infection (affecting 34% and 53% of children, respectively). This is much higher than any estimate for children with infections in the uncontrolled studies (except for one small study^35^), although these children were mostly hospitalized. This underscores that without including a control group, no meaningful interpretation seems possible; it cannot be excluded that reported symptoms may not be caused by the infection. Some studies aimed to investigate the role of prognostic risk factors (such as severity of acute COVID-19) on absolute risks, but these assessments are subject to the same crucial limitations.

The exact relationships and interrelationships of the various factors that influence both the risk of infection and the risk of symptomatic disease need to be understood. These may include age, social factors (e.g., housing conditions, education level of children but also of parents and guardians, family situation), psychological and mental factors (e.g., mental illness or impairment, at least of the children themselves), economic factors (e.g., financial situation and additional financial burden on the family due to the pandemic). This may also include information on the parents as they most likely have serious impact on the children’s risk to be infected and becoming aware of symptoms. The precise influence of such factors on individual risk behaviour is unclear and difficult to model. Moreover, it is difficult to collect very detailed data that would allow such analyses. Of 21 studies, only 6 used control groups, and only two adjusted for some confounders but their results were likely at risk of unmeasured or residual confounding. Hence, for all studies, the risk of confounding was critical. Another level of complexity is the detection of outcomes due to the inconsistently used and non-standardized definition of “long COVID”, but also, and above all, the recording of the outcomes themselves which are often self-reported and subjectively assessed in an unstructured, non-standardized way. A strong association between risk of infection (e.g., being a close contact of a family member) and recognition of an outcome (i.e., recognition of symptoms) can be assumed in this situation. It is conceivable that great concern about “long COVID” leads to more cautious behaviour and more contact restrictions (possibly leading to a lower risk of infection but also higher psychological and mental distress), and to greater attention to symptoms and more frequent contacts with the health care system, increasing the probability of receiving such a diagnosis. Structured prospective data collection with a parallel control group would help to avoid such issues. However, given that most infections are unrecognized, a control group would need to be defined based on a strategy with high sensitivity (e.g., both negative PCR and negative serology).

Overall, the 21 studies had mostly serious risk of bias for the outcome measurement, most studies had missing data for more than 20% of the participants and had a critical risk of selective reporting without clear pre-specification of analyses, without protocols and with unclear definitions of results. We did not consider studies reporting only surrogate outcomes (such as laboratory markers or health care use) unless we found information on pertinent patient-relevant outcomes^24^. Studies were also not eligible if they reported only cumulative incidences of outcomes not allowing to differentiate the acute situation from the longer follow-up and acute symptoms from those persistent over time. We encourage authors of future studies to report more granular details. Previous work has outlined recommendations that can help improve the design, conduct and reporting of such studies^4^. For studies in children, specific issues require special attention. This includes consideration of confounding due to factors not related to the patient but to parents, guardians, and the family that may be associated with both, infection risk and outcome (e.g., educational level, housing situation, financial situation, additional financial burden on the family due to the pandemic). Other critical factors relate to schooling, e.g., remote learning or school closure. All these factors are typically complex and not included in routinely collected data sources, requiring specific and very granular active data collection and careful consideration in the analyses.

### Limitations

Our assessment of risk of bias and study characteristics involve a degree of subjectivism. However, given that many extracted items related to existence of reported information (e.g., availability of study protocols), this left little room for inconsistency. For the risk of bias assessment, vital items were clear and unambiguous, and these alone determined the overall risk of bias assessment (e.g., the critical risk of confounding bias that affects all studies). Here it needs to be highlighted that the applied tool has not been designed for non-interventional studies, but we felt it is the best available choice and it is important to note the underlying logic determining effects is the same for drugs, vaccines, or infections from viruses. Overall, however, the often-limited reporting quality must be considered, so that isolated misinterpretations cannot be ruled out despite our predefined processes. However, this would not change the overall interpretation.

Overall, authors have mostly refrained from causally classifying their results or making exaggerated interpretations of their findings - most study groups have been cautious in their interpretations. While this restraint is commendable, none of the studies communicated the serious limitations of this evidence clearly or called for caution in their conclusions. Inadequate, over-confident interpretation of the findings by media and decision-makers may cause potentially unnecessary fears and worries among parents and children with SARS-CoV-2 infection.

## CONCLUSIONS

Clarifying the frequency and severity of post-COVID syndrome remains an important research aim. The best possible research is needed to clarify the current conundrum.

Children and their families urgently need much more reliable and methodologically robust evidence to address their concerns and improve care.

## Data Availability

All data produced in the present work are contained in the manuscript.

## DECLARATIONS

### Availability of data and materials

All data generated or analyzed during this study are included in this publication and supplement content.

### Ethical approval

Not necessary for this study.

### Authors’ contributions

The authors had full access to all of the data in the study and take responsibility for the integrity of the data and the accuracy of the data analysis.

Concept and design: All authors.

Acquisition, analysis, or interpretation of data: Hirt, Janiaud, Gloy, Schandelmaier, Pereira, Hemkens

Drafting of the manuscript: Hirt, Janiaud, Hemkens

Critical revision of the manuscript for important intellectual content: All authors.

Statistical analysis: Hirt, Janiaud.

Obtained funding: Pereira (Chevening Scholarship Programme).

Administrative, technical, or material support: n/a.

Supervision: Hemkens

