## Supplementary material for "Validity of reported post-acute health outcomes in children with SARS-CoV-2 infection: a systematic review"

Supplement content


- eMethods: Search strategies and documentation
- eFigure1: Literature search and study retrieval process
- eTable1: Expansion of table 2 on main post-acute health outcomes, follow-up length, and details on reporting of methods and symptoms
- eBox1: Reported action recommendations for children based on causal interpretations of their findings
- eTable2: Details and justification on risk of bias assessment (ROBINS-I)
- eReferences

#### eMethods: Search strategies and documentation

##### PubMed (last search: January 22, 2022)

| # | Entry | Hits |
|---|---|---|
| 1 | ((COVID-19) OR (SARS-CoV-2) OR (coronavirus) OR (2019-nCoV)) | 235,644 |
| 2 | ((long-term) OR ("long term") OR ("long haul*") OR (“after recovery”) OR (prolong*) OR (persist*) OR (long-covid*) OR ("long covid*”) OR (post-covid*) OR ("post covid*") OR (post-acute*) OR (“post acute*”)) | 1,754,935 |
| 3 | ((outcome*) OR (symptom*) OR (disease*) OR (illness*)) | 10,045,993 |
| 4 | ((cohort) OR (follow up) OR (longitudinal)) | 3,433,478 |
| 5 | #1 AND #2 AND #3 AND #4 | 3,782 |
| 6 | #5 AND Filters: from 2020/1/1 - 3000/12/12 | 3,726 |

##### Web of Science Core Collection (last search: January 22, 2022)

| # | Entry | Hits |
|---|---|---|
| 1 | TS=((COVID-19) OR (SARS-CoV-2) OR (coronavirus) OR (2019-nCoV)) | 255,865 |
| 2 | TS=((long-term) OR ("long term") OR ("long haul*") OR (“after recovery”) OR (prolong*) OR (persist*) OR (long-covid*) OR ("long covid*”) OR (post-covid*) OR ("post covid*") OR (post-acute*) OR (“post acute*”)) | 2,403,177 |
| 3 | TS=((outcome*) OR (symptom*) OR (disease*) OR (illness*)) | 7,633,107 |
| 4 | TS=((cohort) OR (follow up) OR (longitudinal)) | 2,481,606 |
| 5 | #1 AND #2 AND #3 AND #4 | 2,554 |
| 6 | #5 AND Publication Date from 2021-01-01 to 2021-12-31 | 2,511 |

##### L·OVE (last search: January 25, 2022)

| # | Entry | Hits |
|---|---|---|
| 1 | ((long-term) OR ("long term") OR ("long haul*") OR ("after recovery") OR (prolong*) OR (persist*) OR (long-covid*) OR ("long covid*") OR (post-covid*) OR ("post-covid*") OR (“post-acute*”)) AND ((match*) OR (control*) OR (propensity) OR (seropositive*) OR (seronegativ*)) | 3,633 |
| 2 | Filter “Children & adolescents” | 285 |
| 3 | Manually picking preprints from medRxiv, ResearchSquare, and Social Science Research Network (SSRN) | 75 |

##### EPPI-Centre (last search January 25, 2022)

Separated all field searches in the “Long COVID Segment” using the keywords "children", "adolescents", "paediatric", "pediatric", and "kids" retrieved 18 hits.

**eFigure1: Literature search and study retrieval process**

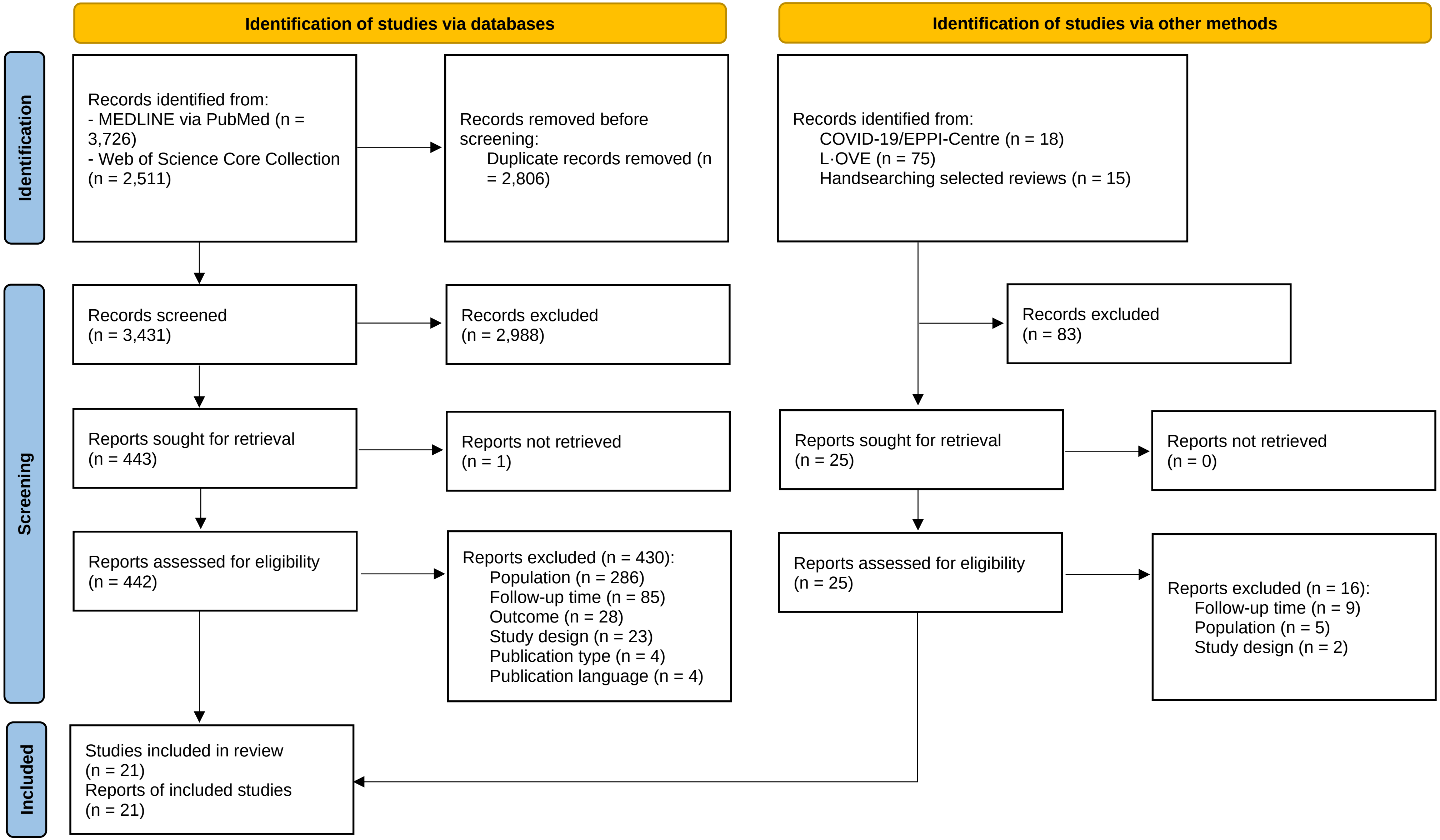

### eTable1: Details on reported post-acute health outcomes (expanded table 2)

| Study | Follow-up (months) * | Outcomes (main outcome) | Number of outcomes (reported/total assessed) | Outcome data collection method | Symptom duration reported | Symptom frequency reported | Symptom trend reported | Symptom severity reported | Subgroup analyses related to main outcome |
|---|---|---|---|---|---|---|---|---|---|
| ***Controlled studies*** | | | | | | | | | |
| Bergia 2021[1] | 4 (infection) | any symptoms; rhinorrhoea; fever; cough; dyspnoea; diarrhoea; vomits; abdominal pain; loss of appetite; headache; anosmia/ageusia; myalgia; asthenia; concentration problems; insomnia; apathy, sad feeling; anxiety; palpitations/tachycardia; dizziness; others | 20/n.r. | Structured questionnaire by phone led by physicians | Yes | - | - | - | Yes ** (gender; hospital stay; days of admission; covid severity; comorbidity; another family member with long COVID; relative long-covid symptoms; age) |
| Blomberg 2021[2] | 6 (infection) | any symptoms; fever; cough; dyspnoea; palpitations; stomach upset; disturbed taste/smell; sleep problems; headache; dizziness; tingling in fingers | 11/11 | Personal interview led by medical staff | - | - | - | - | - |
| Matsubara 2022[3] | 3 (infection) | cardiac symptoms; fatigue; others | 3/n.r. | Routinely collected data (medical records, structured clinical assessment) | - | - | - | - | - |
| Radtke 2021[4] | 6 (infection) | any symptoms; tiredness; difficulty concentrating; increased need for sleep; congested or runny nose; stomachache; chest tightness; headache; sleep disturbances; cough; health status | 11/n.r. | Structured online questionnaire | Yes | - | - | - | - |
| Roessler 2021[5] | 3 (infection) | health outcomes combined; abdominal pain; acute pain; adjustment disorder; anuria/oliguria; anxiety disorder; arthritides; ascites; behavioral symptoms; cachexia; carditis due to viruses; changes in bowel habits; chronic fatigue syndrome; cognitive function impairment; concentration impairment/concentration deficit; cough; covid toe; depression; developmental delay; disorientation; dysgeusia; dyslexia; dysmenorrhea; dysphagia; dyspnea; dysuria; emotional and behavioral disorder; epistaxis; eye pain; facial nerve paralysis; fever; flatulence; gangrene; general symptoms; hair loss; headache; hearing loss/tinnitus; heart failure; heart murmurs; heartburn; hemorrhage; hepatomegaly and splenomegaly; hoarseness; hyperhidrosis; hypotension; impaired balance; joint pain; loss of appetite; lymphadenopathy; malaise/fatigue/exhaustion; memory impairment; meningism; mood disorder; mood disorder; movement disorders; myalgia; myocardial infarction; myocarditis; nausea; neurasthenia; neurological manifestation of post-covid; obsessive-compulsive disorder; oedema; other cardiac arrhythmias; other coordination disorders/ataxia; other symptoms of the urinary system; pain, not elsewhere classified; paresis; paresthesia of skin; pathological findings from male genital tract; pathological lung findings; pathological reflexes; pericarditis; polyuria; post-covid; pulmonary embolism; rash; respiratory insufficiency; seizures; sensation and perception disorder; shock; sinus vein thrombosis; sleep disorders; somatization disorder; somnolence; sopor/coma; speech and language disorders; stroke; subcutaneous nodules; syncope; tetany; throat/chest pain; thrombosis; urethral discharge; urinary retention; vertigo; visual disturbances; weight gain/loss, eating disorders | 97 (grouped)/97 | Routinely collected data (administrative claims; unclear how symptoms were assessed) | - | - | - | - | Yes (severity of COVID-19 (hospitalized, intensive care unit, outpatient) stratified by age group; diagnosis/symptom complex stratified by age group) |
| Stephenson 2021[6] | 3 (infection) | any symptoms; fever; chills; persistent cough; tiredness; shortness of breath; loss of smell; unusually hoarse voice; unusual chest pain; unusual abdominal pain; diarrhoea; headaches; confusion, disorientation or drowsiness; unusual eye-soreness; skipping meals; dizziness or light-headedness; sore throat; unusual strong muscle pains; earache or ringing in ears; raised welts on skin or swelling; red/purple sores/blisters on feet; other; quality of life/functioning; fatigue; mental health and wellbeing | 25/25 | Structured online questionnaire | - | - | - | - | Yes (age group) |
| ***Uncontrolled studies*** | | | | | | | | | |
| Asadi-Pooya 2021[7] | 8 (recovery) | any symptoms; muscle weakness; muscle pain; joint pain; fatigue; sleep difficulty; anxiety; depression; shortness of breath; chest pain; palpitation; cough; excess septum; decreased sense of smell; decreased sense of taste; sore throat; headache; dizziness; concentration difficulty; excess sweating; exercise difficulty; walking difficulty; diarrhoea; abdominal pain/stomachache; loss of appetite; skin lesions; other; chronic medical illness/problem | 28/28 | Structured questionnaire by phone (unclear who led by) | - | Yes | - | Yes | Yes (sex; age; length of hospital stay; symptoms at presentation (fever, respiratory distress, cough, muscle pain, diarrhoea, intensive care unit admission) |
| Bottino 2021[8] | 2 (recovery) | respiratory symptoms | 1/n.r. | Clinical visit (unclear who led by and how | - | - | - | - | - |

| | | | | | | | | | |
|---|---|---|---|---|---|---|---|---|---|
| | | | | symptoms were assessed) | | | | | |
| Capone 2021[9] | 6 (infection) | fatigue | 1/n.r. | Clinical visit (unclear who led by and how symptoms were assessed) | - | - | - | - | - |
| Chowdhury 2021[10] | 5 (recovery) | any symptoms; lethargy; cough; chest discomfort and pain; fatigue; breathlessness on activity; anxiety, lack of concentration, and occasional amnesia; headache; fever (persisting fever); joint pain; mild body ache; chill; enteric fever; back pain; type 2 diabetes mellitus (following covid-19); hypertension (following covid-19) | 16/n.r. | Phone call (unclear who led by and which symptoms were assessed) | Yes | - | - | - | - |
| Denina 2020[11] | 4 (recovery) | any symptoms | 1/n.r. | Phone call (unclear who led by and which symptoms were assessed) | - | - | - | - | - |
| Isoldi 2021[12] | 6 (infection) | any symptoms | 1/n.r. | Clinical visit (unclear who led by and how symptoms were assessed) | Yes | - | - | - | - |
| Kahn 2022[13] | 2 (infection) | any symptoms; fatigue; muscle/joint weakness/pain; skin manifestations; gastrointestinal symptoms; reduced exercise capacity; psychiatric or neuropsychiatric problems; headache; others | 9/n.r. | Routinely collected data (registry; structured clinical assessment) | - | - | - | - | - |
| Matteudi 2021[14] | 11.5 (recovery) | any symptoms; late-onset symptoms; recovery from symptoms; asthenia; learning difficulties; headache | 6/n.r. | Phone call led by a paediatric team (unclear which symptoms were assessed) | - | - | Yes | - | Yes (age group; symptomatic/asymptomatic during the acute phase; hospitalization) |
| Mei 2021[15] | 5 (recovery) | any symptoms; shortness of breath; cough/sputum; pharyngitis/foreign body feeling; dyspnoea; pulmonary fibrosis; lung damage; bronchitis; copd; haemoptysis; chest pain/tightness; palpitation; cardiac disease; tachycardia; angina pectoris; heart attack; insomnia; joint pain/back pain/lumbago; fatigue; headache/dizziness/poor memory; change of taste and smell; myalgia; impaired vision; leg numbness/finger stiffness; neuralgia; paralysis; tinnitus; confusion; coma; cerebral infarction; hair loss; bitter/dryness in mouth; high blood sugar; diabetes; gastrointestinal complaints/poor appetite; diarrhoea; constipation; emesis; hidrosis; erythron; allergy; hepatic insufficiency; enema; antiadoncus; hypertension; kidney insufficiency; reduction of physical strength; dryness/excessive secretion | 1/48 | Case reports, medical records, self-reports (unclear who led by and how symptoms were assessed) | - | - | - | - | - |
| Osmanov 2021[16] | 8.5 (recovery) | any symptoms; fatigue; nasal congestion/rhinorrhoea; insomnia; disturbed smell; headache; disturbed taste; hyperhidrosis; persistent cough; hypersomnia; poor appetite; skin rash; diarrhea; stomach/abdominal pain; problems seeing/blurred vision; hair loss; dizziness/light headedness; joint pain or swelling; variations in heart rate; constipation; loss of smell; difficulty breathing/chest tightness; palpitations; feeling nauseous; chest pain; persistent muscle pain; problems with balance; urination problems; vomiting; confusion/lack of concentration; pain on breathing; cannot fully move or control movement; tremor/shakiness; bleeding; changes in menstruation; loss of taste; tingling feeling/“pins and needles“; weight loss; problems swallowing or chewing; bilateral conjunctivitis; seizures/fits; lumps or rashes (purple/pink) on toes; problems speaking or communicating; fainting/blackouts | 44/44 | Structured questionnaire by phone led by medical students | Yes | Yes | Yes | - | Yes (age group; sex; neurological conditions; allergic diseases; gastrointestinal problems; excessive weight and obesity; COVID severity) |
| Patnaik 2021[17] | 3.5 (recovery) | any symptoms | 1/n.r. | Clinical visit (unclear who led by and how | - | - | - | - | - |

| | | | | | | | | | |
|---|---|---|---|---|---|---|---|---|---|
| | | | | symptoms were assessed) | | | | | |
| Penner 2021[18] | 6 (infection) | gastrointestinal symptoms; persistent abdominal pain; persistent diarrhoea; new-onset nausea and vomiting; new-onset diarrhoea; dysphonia; anosmia or dysgeusia; dysphagia; rashes | 9/9 | Routinely collected data (medical records; structured clinical assessment) | Yes | - | Yes | - | - |
| Rusetsky 2021[19] | 2 (recovery) | olfactory disorder | 1/1 | Structured questionnaire by phone led by the investigators | Yes | - | - | Yes | Yes (age group; gender) |
| Say 2021[20] | 4.5 (infection) | any symptoms; fever >38; sore throat; cough; runny nose; shortness of breath; loss of taste; loss of smell; poor appetite; vomiting; low energy or tiredness; headaches; muscle aches and pains; abdominal pain; diarrhoea; other; shortness of breath; fatigue; rash; fever; abdominal pain; conjunctivitis; wellbeing; immunisation reaction | 24/24 | Clinical visit using a structured clinical assessment (unclear who led by) | Yes | - | - | - | - |
| Sterky 2021[21] | 7 (infection) | any symptoms; fever; elevated pulse; palpitations; difficulties breathing; headache; fatigue; increased need of sleep; decreased activity level; decreased physical strength; concentration difficulties; reduced/changed taste; loss of appetite; affected memory; difficulties managing school; depressive symptoms; recurrent body pains; other diagnosed illness | 18/18 | Structured questionnaire by phone (unclear who conducted) | - | - | - | Yes | Yes (age group; symptoms at onset; treatment received; days of hospitalization; c-reactive protein; days since discharge; chronic illness) |

* Follow-up started at detection of infection, onset of symptoms, COVID-19 diagnosis and/or hospital admission (described as infection) or at recovery from the acute illness and/or hospital discharge (described as recovery). Median follow-up length if reported by the authors, we converted weeks and days to months[3,8,15,16,21] (4 weeks/30 days = 1 month); if the median follow-up was not available, we assumed the half of the range[2,7,14,17,20] or the interquartile range[11] to be the median; if both median and range were not available, the fixed follow-up length as reported by the authors has been chosen for our analysis[1,4-6,9,12,18,19]; one study reported outcome data for multiple follow-up lengths between 1 and 5 months, here, we report on 5 months follow-up[10]; one study reported outcome data for multiple follow-up lengths between 2 and 6 months, here, we report on 6 months follow-up[13].

** Data for subgroups only reported for children with SARS-CoV-2 infection.

Abbreviations: n.r. = not reported.

**eBox1: Reported action recommendations for children based on causal interpretations of their findings**

- "Considering the millions of young people infected during the ongoing pandemic, our findings are a strong impetus for comprehensive infection control and population-wide mass vaccination." [2]
- "To the extent that these results reflect a higher long-run morbidity related to SARS-CoV-2 infections, they may indicate an important public health challenge that should be considered in discussions about adequate preventive measures." [5]

**eTable2: Details and justification on risk of bias assessment (ROBINS-I)**

| | Asadi-Pooya 2021 | Bergia 2021 | Blomberg 2021 | Bottino 2021 | Capone 2021 | Chowdhury 2021 | Denina 2020 | Isoldi 2021 | Kahn 2022 | Matsubara 2022 | Matteudi 2021 | Mei 2021 | Osmanov 2021 | Patnaik 2021 | Penner 2021 | Radtke 2021 | Roessler 2021 | Rusetsky 2021 | Say 2021 | Stephenson 2021 | Sterky 2021 |
|---|---|---|---|---|---|---|---|---|---|---|---|---|---|---|---|---|---|---|---|---|---|
| **Bias due to confounding** | C | C | C | C | C | C | C | C | C | C | C | C | C | C | C | C | C | C | C | C | C |
| **Bias due to selection of participants** | C | C | C | C | C | C | C | S | C | C | C | S | C | C | S | S | L | C | C | L | C |
| **Bias in classification of infection status/exposure** | L | S | M | L | L | L | L | L | L | S | L | L | L | L | L | M | S | L | NI | M | L |
| **Bias due to missing data** | S | S | S | L | S | S | S | S | S | S | S | M | S | S | L | S | L | L | L | S | L |
| **Bias in measurement of outcomes** | S | S | S | NI | NI | S | S | NI | M | M | S | NI | S | NI | M | S | S | S | M | S | S |
| **Bias in selection of the reported result** | C | C | C | M | C | C | C | C | C | C | C | C | M | C | C | C | M | M | C | C | M |

C: Critical risk of bias; S: Serious risk of bias; M: Moderate risk of bias; L: Low risk of bias; NI: No information to assess respective risk of bias.

#### BIAS DUE TO CONFOUNDING

All uncontrolled studies, by design, are at a critical risk of confounding bias.

**Roessler 2021** and **Stephenson 2021** used a matched case-control design. However, none of them adjusted the analysis for potential confounders such as housing and family setting and socioeconomic status that we assume to be likely associated with the risk of being exposed to SARS-CoV-2 and of reporting long COVID symptoms.

**Radtke 2021** used a control group but did not make any formal comparison between the cases and controls nor mentioned the issue of confounders.

**Blomberg 2021** used a control group but did not make any formal comparison between the pediatric cases and control nor mentioned the issue of confounders.

Although **Bergia 2021** and **Matsubara 2022** used a control group and alluded to confounding in the discussion and **Bergia 2021** mentioned that findings may be affected by residual confounding, and **Matsubara 2022** acknowledged specific non-adjusted confounders, both studies did not make any formal comparison between the pediatric cases and control.

#### BIAS DUE TO SELECTION OF PARTICIPANTS

Studies were deemed as having critical risk of bias if they included only hospitalized participants and/or participants who were aware of their COVID-19 status and/or children with multisystem inflammatory syndrome and were more likely to participate to the study if they had persistent symptoms due to self-selection.

**Blomberg 2021** included outpatient and hospitalized patients; however the selection of the infection-negative was made from the same households as the cases and some cases were defined on serology with no clear indication if the timing of the infection. It was thus assessed at critical risk of bias.

**Isoldi 2021**, **Mei 2021,** and **Penner 2021** were deemed as having serious risk of bias as they included hospitalized participants but followed all participants until the end and there was no risk of self-selection.

**Roessler 2021** and **Stephenson 2021** were deemed as having low risk of bias as cases and controls were selected from public health databases with hospitalized, non-hospitalized, symptomatic, and asymptomatic patients. In addition, in **Roessler 2021** outcome data were collected from electronic health records and in **Stephenson 2021** outcome data were collected through an online questionnaire with similar response rates in both groups; thus both studies were deemed at low risk of self-selection.

**BIAS IN CLASSIFICATION OF INFECTION STATUS/EXPOSURE**

Uncontrolled studies were assessed as having low risk of bias, if the infection status classification was based on RT-PCR, antigen or serology testing during the acute phase of COVID-19.

**Say 2021** did not provide information on the type of test/diagnosis and could not be assessed.

**Radtke 2021** used serology for infection status classification with no clear indication of the timing of the infection for the positive SARS-CoV-2 infection status.

**Blomberg 2021** defined positive SARS-CoV-2 infection status using RT-PCR test and in some cases serology testing while all negative SARS-CoV-2 infection status was defined using serology.

**Stephenson 2021** SARS-CoV-2 infection status was defined using RT-PCR; however, it cannot be excluded that participants identified as SARS-CoV-2 negative had been infected in the past.

**Roessler 2021** did not provide information on the type of testing, but positive SARS-CoV-2 infection status was defined as documented COVID-19 diagnosis with confirmed laboratory virus detection in the health insurance database while negative SARS-CoV-2 infection status was defined as lack of a documented COVID-19 diagnosis regardless of there was a record of laboratory virus detection.

**Matsubara 2022** and **Bergia 2021** defined positive SARS-CoV-2 infection status using PCR test or serology, but controls were defined as pre-pandemic (**Matsubara 2022**) or as having no COVID-19 symptoms (**Matsubara 2022 and Bergia 2021),** testing was not required for controls.

**BIAS DUE TO MISSING DATA**

**Bottino 2021, Penner 2021, Rusetsky 2021, Say 2021,** and **Sterky 2021** were assessed as having low risk of bias as authors reported no or minimal loss to follow-up (≤ 20%). **Roessler 2021** was also assessed as having low risk of bias as data were collected using systematically routinely collected data.

**Mei 2021** was assessed as having a moderate risk of bias as, although report following all eligible participants, data were collected from multiple data sources without further details (i.e., case reports, medical records, and self-reports).

**Matsubara 2022** was assessed as having a serious risk of bias with >20% loss to follow-up in children with positive SARS-CoV-2 infection status and unclear loss to follow-up in controls.

All other studies were assessed as having a serious risk of bias and had >20% loss to follow-up except for **Asadi-Pooya 2021** and **Denina 2020** who report unclear information regarding missing data.

**BIAS IN MEASUREMENT OF OUTCOMES**

Studies deemed as having serious risk of bias if the outcomes were reported by the participants who had knowledge of their COVID-19 status; thus, cases were more likely to report subjective symptoms.

**Roessler 2021** deemed as having serious risk of bias since they used routinely collected data but did not provide sufficient information on how the symptoms were assessed.

**Say 2021** deemed as having moderate risk of bias since the outcomes were assessed during a clinical visit using structure clinical assessments but the outcome assessor was likely to be more attentive as aware of the COVID-19 status of participants.

**Matsubara 2022, Penner 2021,** and **Kahn 2022** deemed as having moderate risk of bias since they used routinely collected data based on structured clinical assessment.

For **Isoldi 2021, Bottino 2021, Patnaik 2021, Mei 2021,** and **Capone 2021**, no detailed information is provided for the outcome measurement and the risk of bias could not be assessed.

**BIAS IN SELECTION OF THE REPORTED RESULT**

Studies deemed as having critical risk of bias if they did not provide a protocol, did not clearly define their outcomes in the method section, did not provide the questionnaire as supplement and/or clearly did not report all outcomes.

Studies deemed as having moderate risk of bias if they did not provide a protocol (or the protocol was not freely available **Osmanov 2021**) but either defined their outcome in the methods section (**Bottino 2021** and **Rusetsky 2021**) or provided the questionnaire used as supplement (**Sterky 2021, Roessler 2021,** and **Osmanov 2021**) and reported on all outcomes.
